# National trends of unique-PCV20 / PCV21 serotypes, common PCV20-PCV21 and non-vaccine serotypes causing invasive pneumococcal disease in adults in Spain during the period 2009-2026

**DOI:** 10.64898/2026.09.22.26363691

**Authors:** Elsa García-González, Julio Sempere, Mirella Llamosí, Covadonga Pérez-García, Aída Úbeda, Andrea Sánchez-Honrado, Loreine Agulló, Carmen Ardanuy, Laura Pérez-Fernández, Alain Ocampo-Sosa, Elisa Ramos-Sevillano, Jeremy S Brown, Mirian Domenech, Jose Yuste

**Author notes:** **Correspondence to:** Mirian Domenech PhD,; Jose Yuste PhD,. National Center for Microbiology, Instituto de Salud Carlos III, Ctra Majadahonda-Pozuelo Km 2, 28220 Madrid, Spain. EGG and JS contributed equally to this work.

## Abstract

**Objectives:** To evaluate the epidemiological evolution of unique pneumococcal serotypes covered by either PCV20 or PCV21, common serotypes by both vaccines and non-vaccine serotypes in adults.

**Methods:** We characterized all national IPD clinical isolates from January 2009 to June 2026.

**Results:** The highest burden of IPD was caused by serotypes shared between PCV20 and PCV21 with serotypes 3 and 8 the most prevalent in adults of all ages. During the pre-pandemic period, IPD caused by unique-PCV20 serotypes declined probably due to herd protection by paediatric vaccination with PCVs. In contrast, unique-PCV21 serotypes increased in incidence accounting for more cases than unique-PCV20 serotypes. However, after the COVID-19 pandemic, a different pattern was observed with unique-PCV20 serotypes increasing in incidence due a marked rise in IPD caused by serotypes 4 and 14 that has progressively reduced the gap in disease coverage. In adults aged 18-64 years, the fraction of disease that could be prevented with PCV20/PCV21 changed from 5% in favour of PCV21 (years 2022/2023) to 15% in favour of PCV20 (year 2026) whereas in adults ≥65 years the vaccine contribution declined from 20% in favour of PCV21 (year 2022) to 8% in 2026.

**Conclusions:** IPD trends in adults have changed after the COVID-19 pandemic, especially in recent years. Our study emphasizes the rapid re-emergence of infections caused by traditional PCV7 serotypes especially serotypes 4 and 14 and reinforce the idea that the use of broader PCVs must be accompanied by an increase in the uptake rates in adults of all ages.

## Introduction

The recent licensure of two new broader pneumococcal conjugate vaccines (PCVs) in adults, termed PCV20 or PCV21, benefits the prevention of invasive pneumococcal disease (IPD). The approval of these new PCVs raises the question of which vaccine should be recommended by the public health authorities because their composition differ substantially, as they share common serotypes that are present in both PCVs, but they also contain unique serotypes that differ in each formulation. The PCV20 vaccine covers all 13 serotypes included in the PCV13 vaccine plus seven additional serotypes. In contrast, the PCV21 vaccine was originally targeted for adults, and the composition differs from PCV20 because this vaccine lacks several serotypes present in PCV13 and added 11 serotypes that are only covered by PCV21. Hence, introduction of these vaccines, presents a challenge for the epidemiological evaluation of prevalent serotypes causing IPD in adults. In this context, national microbiology reference laboratories play an essential role in the surveillance of vaccine effectiveness and the impact of these new PCVs in the real world, monitoring changes in serotype distribution, in both target and non-target populations (1, 2). In Spain, PCV13 was introduced in 2016 in the national paediatric immunization program (2+1 schedule) with high uptake rates (>95% even during the COVID-19 pandemic) although it was previously used in the private market between 2010-2015 with high coverage rates up to 82% (3-5).

Epidemiological surveillance studies in Spain have demonstrated that the paediatric use of PCV7 followed by PCV13 has significantly reduced the burden of disease by vaccine-serotypes not only in children (direct effect) but also in adults (herd protection) (3, 4, 6). However, hospitalization rates in adults due to pneumococcal infections are still very high despite the use of PCVs, and this is a critical aspect that deserves attention, because pneumococcal disease is still a major cause of morbidity and mortality, especially in older adults, that are associated with high medical costs of more than EUR 359 million per year (7, 8). The reason for these high incidence rates of pneumococcal disease in adults is complex, but several factors are major contributors, such as low vaccination uptake in adults, immunosenescence leading to reduced immunogenicity, and the rise of certain lineages with increased potential to avoid the host immune response and cause disease (6, 9-13).

The use of PCVs in Spanish immunocompetent adults has been heterogeneous with great variations within the country. PCV13 was introduced in 2016 in 6 of 17 Spanish regions, whereas 11 regions were still using the 23-valent polysaccharide vaccine (PPSV23). In 2024 all Spanish regions changed to PCV20 and in 2026, 3 of 17 regions have switched to PCV21, although one common aspect in all the country is the low uptake rate achieved in adults from 8.48% in 2018 to 35.51% in 2025 (data supplied from Spanish Ministry of Health; https://pestadistico.inteligenciadegestion.sanidad.gob.es/publicoSNS/S/sivamin). Hence, this vaccination coverage rate does not reflect true PCV uptake in adults, as it aggregates data on adults who have received at least one pneumococcal vaccine dose at any point in their lifetime.

In this study, we have included the epidemiological distribution of pneumococcal serotypes causing IPD in adults of different group ages before and after the COVID-19 pandemic, including the period January 2009 to June 2026. To assess the potential contribution of PCV20 / PCV21 we have analysed the distribution of common serotypes present in both vaccines, unique-PCV20 serotypes (only present in PCV20 but not in PCV21), unique-PCV21 (only present in PCV21 but not in PCV20) and non-vaccine serotypes (not included in the formulation of PCV20 and PCV21 vaccines).

## Methods

### Study design

This is a national longitudinal surveillance study that included all IPD isolates (37,439) from hospitalized adults ≥ 18 years reported to the Spanish Pneumococcal Reference Laboratory (SPRL) during the period January 1^st^, 2009, to June 30^th^, 2026.

The SPRL is a member of the IBD-labnet funded by the European Center for Disease Control (ECDC) and notifies to ECDC all these IPD cases in Spain, which cover up to 80% of the national level according to estimates by the National Center for Epidemiology. Moreover, since 2018 the SPRL also notifies the IPD cases to the Invasive Respiratory Infection Surveillance Network (IRIS) (1, 2). Serotyping was performed using Quellung reaction, dot blot assay using specific antisera, and/or PCR- capsular sequence typing (4, 6). In some strains, several of these assays were performed simultaneously to identify the serotype correctly.

### Data processing

The epidemiological year considered in the manuscript is from January to December (2009-2025), except data of 2026, which only contained the first semester. Serotypes were grouped into four different categories: common-PCV20-21 (containing serotypes shared in both vaccines PCV20 and PCV21), unique-PCV20 serotypes (serotypes only present in PCV20 but not in PCV21), unique-PCV21 (serotypes only present in PCV21 but not in PCV20) and non-vaccine (serotypes that are not present in PCV20 and PCV21). Serotypes 15B and 15C were classified as common-PCV20-21 types due to cross-reactivity as they have been reported as a single entity in recent epidemiological studies (14-18). Serotypes included in each category described above are indicated as follows:

- Common-PCV20-21 serotypes (3, 6A, 7F, 8, 10A, 11A, 12F, 15B/C, 19A, 22F, 33F).
- Unique-PCV20 serotypes (1, 4, 5, 6B, 9V, 14, 18C, 19F, 23F).
- Unique-PCV21 serotypes (9N, 15A, 16F, 17F, 20, 23A, 23B, 24F, 31, 35B).
- Non-vaccine serotypes (serotypes not included in any of the categories above).

IPD evolution was analysed for different age groups covering the adult population (18-64 years, 18–49 years, 50-64 years, 65-74 years, 75-84 years, ≥85 years and ≥65 years). Three reference years were selected a priori to capture key epidemiological transitions: 2009 (pre-PCV13 introduction in the national immunization program), 2019 (last complete pre-COVID-19 pandemic year), and 2025 (last complete post-pandemic epidemiological year with available data). These anchor points allow direct comparison of the pre-vaccine, pre-pandemic, and current post-pandemic epidemiological landscape. Incidence rate ratios (IRR) were calculated comparing different periods.

Potential serotype coverage was defined as the proportion of preventable IPD by PCV20 and PCV21 in adults of age groups 18-64, 18-49, 50-64, ≥65, 65-74 and ≥75 years for the period 2019-2026.

### Statistical analysis

The incidence was calculated as the number of IPD episodes per 100,000 population and year using population data from the Spanish National Statistical Institute as denominator. The corrected incidence was calculated by applying the population capture of 80% to the denominator. Comparison of different periods was evaluated by calculating the incidence rate ratio (IRR) using Poisson regression models. Statistical analyses were performed using STATA v.14.

## Results

### Evolution of IPD in adult population

In all age-groups, common-PCV20-21 serotypes were responsible for the larger burden of IPD accounting for more cases than unique-PCV20/21 or non-vaccine serotypes, and this pattern was consistent across the pre-pandemic period (2009-2019), the pandemic period (2020-2021) and the reopening/post-COVID period (2022-2026) (**Figures 1 and 2**). Patterns show that despite an initial reduction of common-PCV20-21 serotypes in the first years of PCV13 introduction, incidence resurged rapidly in the late vaccine period, and again after the COVID-19 pandemic, showing almost no difference in incidence for all adults when comparing the last full epidemiological year (2025) versus the pre-pandemic period (2009) (IRR, 1.05; 95% CI, 0.97–1.13 for the age group ≥65 years old and IRR, 1.04; 95% CI, 0.96–1.12 for adults aged 18–64 years old, **Figure 3 and Supplementary Table 1**).

**Fig 1:**
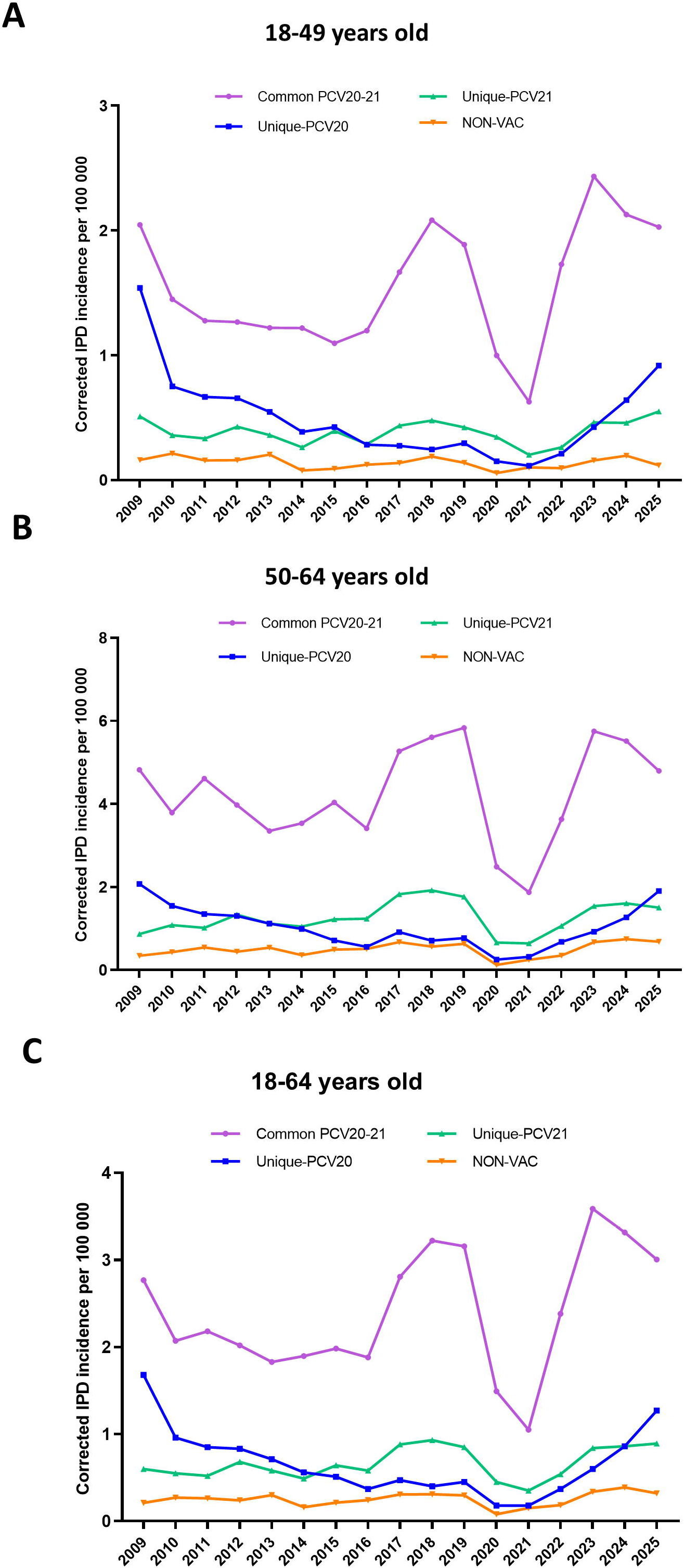
Evolution of IPD corrected incidence in young adults of different age groups during 2009-2025. Serotypes are shown as common PCV20/21 (purple colour; serotypes present in both PCV20 and PCV21) vs unique-PCV20 (blue colour; serotypes only present in PCV20 but not in PCV21) vs unique-PCV21 (green colour; serotypes present in PCV21 but not in PCV20) vs non-VAC (orange colour; serotypes that are not present in PCV20 or PCV21). Adults aged 18-49 years (A), 50-64 years (B), and all young adults (18-64) are represented.

**Fig 2:**
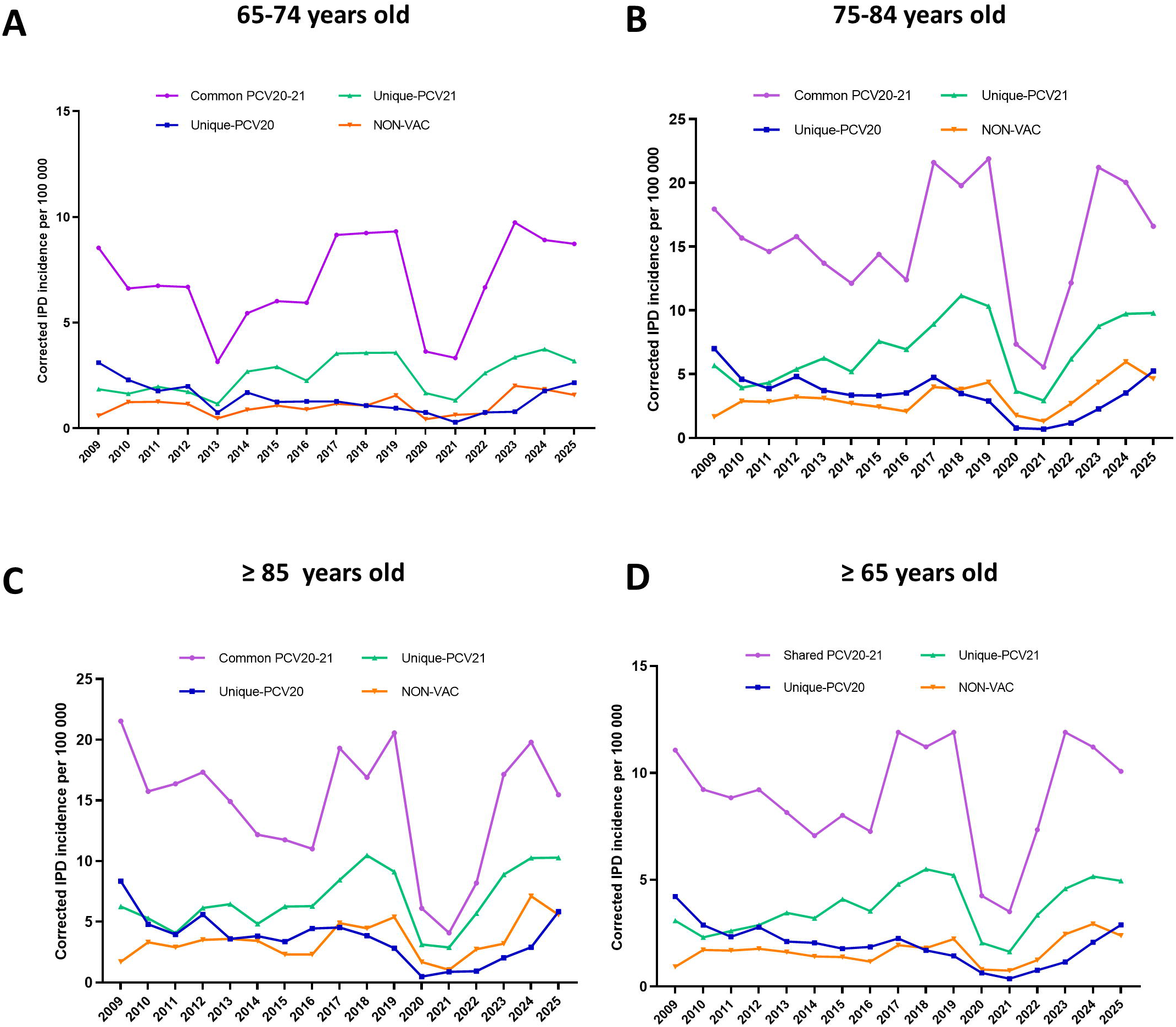
Evolution of IPD corrected incidence in adults ≥ 65 years of age during 2009-2025. Serotypes are shown as shared PCV20/21 (purple colour; serotypes present in both PCV20 and PCV21) vs unique-PCV20 (blue colour; serotypes only present in PCV20 but not in PCV21) vs unique-PCV21 (green colour; serotypes present in PCV21 but not in PCV20) vs non-VAC (orange colour; serotypes that are not present in PCV20 or PCV21). Adults aged 65-74 years (A), 75-84 years (B), ≥85 years old (C), and all older adults (≥65) (D) are represented.

**Fig 3:**
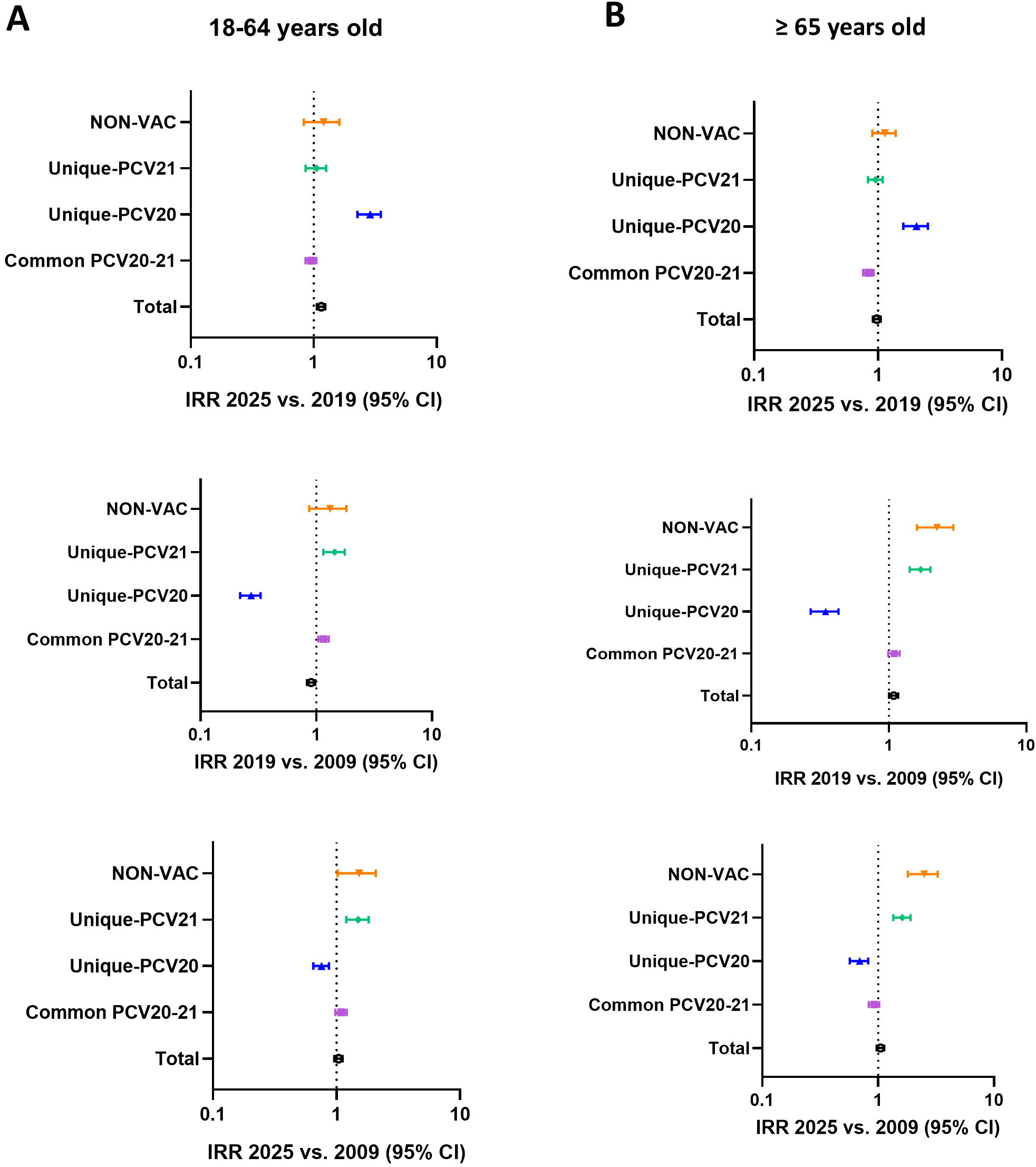
Forest plot of the IRRs comparing the last full epidemiological year (2025) versus the late vaccine period (2019) and pre-PCV13 period (2009). All young adults aged 18-64 years (A) and older adults aged ≥65 (B) are represented. An IRR of more than 1 indicates a higher incidence rate in the comparator or exposed group (period 1) than in the reference group (period 2), whereas an IRR of less than 1 (but >0) indicates a lower incidence rate in the comparator or exposed group (period 1) than in the reference group (period 2). Error bars represent 95% CI.

In young adults, unique-PCV20 serotypes were more frequent than unique-PCV21 serotypes before PCV13 was introduced in Spain (2009) and during the early vaccine period (2010-15) (**Figure 1**). Unique-PCV20 incidence then declined significantly, likely due to indirect immune protection (IRR 2019 vs. 2009, 0.27; 95% CI, 0.22–0.33, **Figure 3 and Supplementary Table 1**). However, following the COVID-19 pandemic, unique-PCV20 incidence rose sharply, surpassing unique-PCV21 incidence and nearly returning to pre-PCV13 levels (IRR 2025 vs. 2019, 2.83; 95% CI, 2.27–3.52 and IRR 2025 vs. 2009, 0.75; 95% CI, 0.65–0.87, **Figure 3 and Supplementary Table 1**). In contrast, unique-PCV21 serotypes became more frequent than unique-PCV20 serotypes during the late PCV13 period, reflecting a steady increase since PCV13 introduction (IRR 2019 vs. 2009, 1.42; 95% CI, 1.15–1.76), a trend that continued into the first post-COVID-19 years (**Figures 1, 3, and Supplementary Table 1**). However, in the most recent epidemiological years, unique-PCV21 incidence remained stable (IRR 2025 vs. 2019, 1.04; 95% CI, 0.86–1.26) and cases were exceeded by unique-PCV20 serotypes in all young adult populations (**Figures 1, 3, and Supplementary Table 1**). Non-vaccine serotypes incidence remained low during all the study period (**Figure 1**).

In older adults, unique-PCV21 showed higher incidence rates than unique-PCV20 serotypes for all age groups in most of the study period (**Figure 2**). Since PCV13 introduction in the paediatric population, unique-PCV20 serotypes showed a decline pattern due to herd protection, reaching the lowest incidence rates during the pre-pandemic and COVID-19 periods (IRR 2019 vs. 2009, 0.34; 95% CI, 0.27–0.43, **Figure 3 and Supplementary Table** 1). In parallel, unique-PCV21 serotypes incidence increased (IRR 2019 vs. 2009, 1.69; 95% CI, 1.42–2.01, **Figure 3 and Supplementary Table 1**). However, during the late post-COVID-19 period (2024/2025) unique-PCV21 serotypes have reached a plateau (IRR 2025 vs. 2019, 0.95; 95% CI, 0.83–1.09) whereas unique-PCV20 serotypes show a continuous rise reducing the differences between PCV20 and PCV21 (IRR 2025 vs. 2019, 2.01; 95% CI, 1.60–2.53, **Figures 2, 3, and Supplementary Table 1**). In contrast to younger adults, non-vaccine serotype incidence rates increased during the full epidemiological period reported in the present study (IRR 2025 vs. 2009, 2.43; 95% CI, 1.80–3.27) but have remained stable since the pre-COVID-19 period (IRR 2025 vs. 2019, 1.12; 95% CI, 0.90–1.39, **Figures 2, 3, and Supplementary Table 1**).

In the first semester of 2026, unique-PCV20 serotypes were more frequently found than unique-PCV21 serotypes in all age groups affecting young adults (18-64 years) (**Figure 4**). In older adults (≥ 65 years), unique-PCV21 serotypes caused higher numbers of IPD cases than unique-PCV20 serotypes (**Figure 4**).

**Fig 4:**
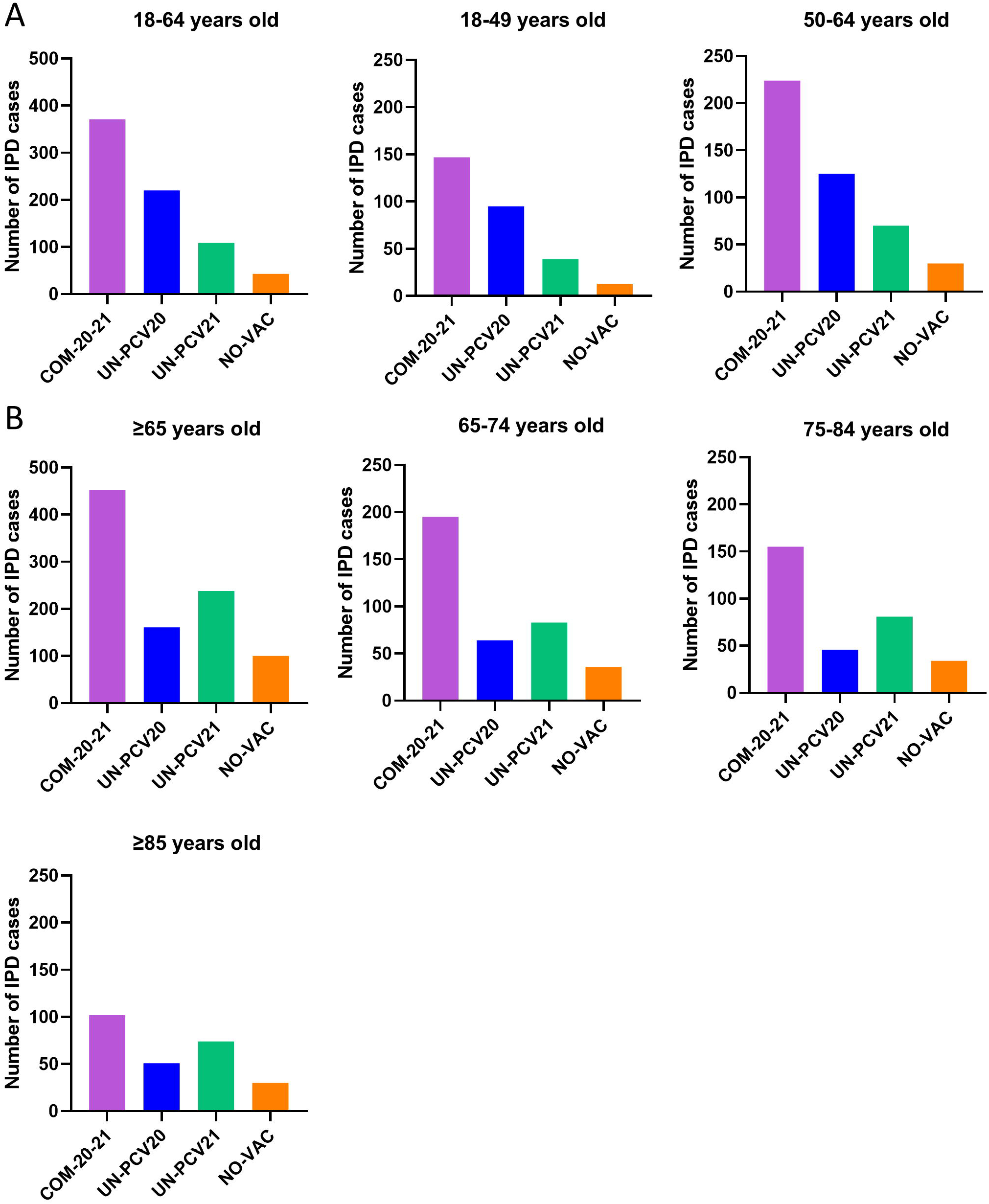
Number of IPD cases caused by common serotypes in PCV20-21, unique- PCV20, unique-PCV21 and NON-Vaccine serotypes during the first semester of 2026 in adults of different age groups. All young adults aged 18-64 years (A) and older adults aged ≥65 (B) are represented. Serotypes are shown as shared PCV20/21 (purple colour; serotypes present in both PCV20 and PCV21) vs unique-PCV20 (blue colour; serotypes only present in PCV20 but not in PCV21) vs unique-PCV21 (green colour; serotypes present in PCV21 but not in PCV20) vs non-VAC (orange colour; serotypes that are not present in PCV20 or PCV21).

### Contribution of PCV20/PCV21 in the prevention of adult IPD

To evaluate the benefit of PCV21 vs PCV20 in the prevention of IPD in adults, we evaluated the fraction of disease that can be prevented by each vaccine during the epidemiological period of 2019-2026 in different age groups (**Figure 5**). We analysed the proportion of IPD cases caused by unique serotypes contained in each vaccine during 2019-2026, and we calculated the difference in the potential fraction of preventable disease between unique-PCV21 serotypes vs unique-PCV20 serotypes **(Figures 5A-B**). In young adults, the fraction of disease that could be prevented with both vaccines swapped from around 12% in favour of PCV21 (years 2019-2021) to up to 19% in favour of PCV20 during 2026 (**Figure 5A**). Hence, the benefit for PCV20 vs PCV21 in 2026 was 15% for the age group 18-64 years, 19% for 18-49 years and 12% for 50-64 years old (**Figure 5A**).

**Fig 5:**
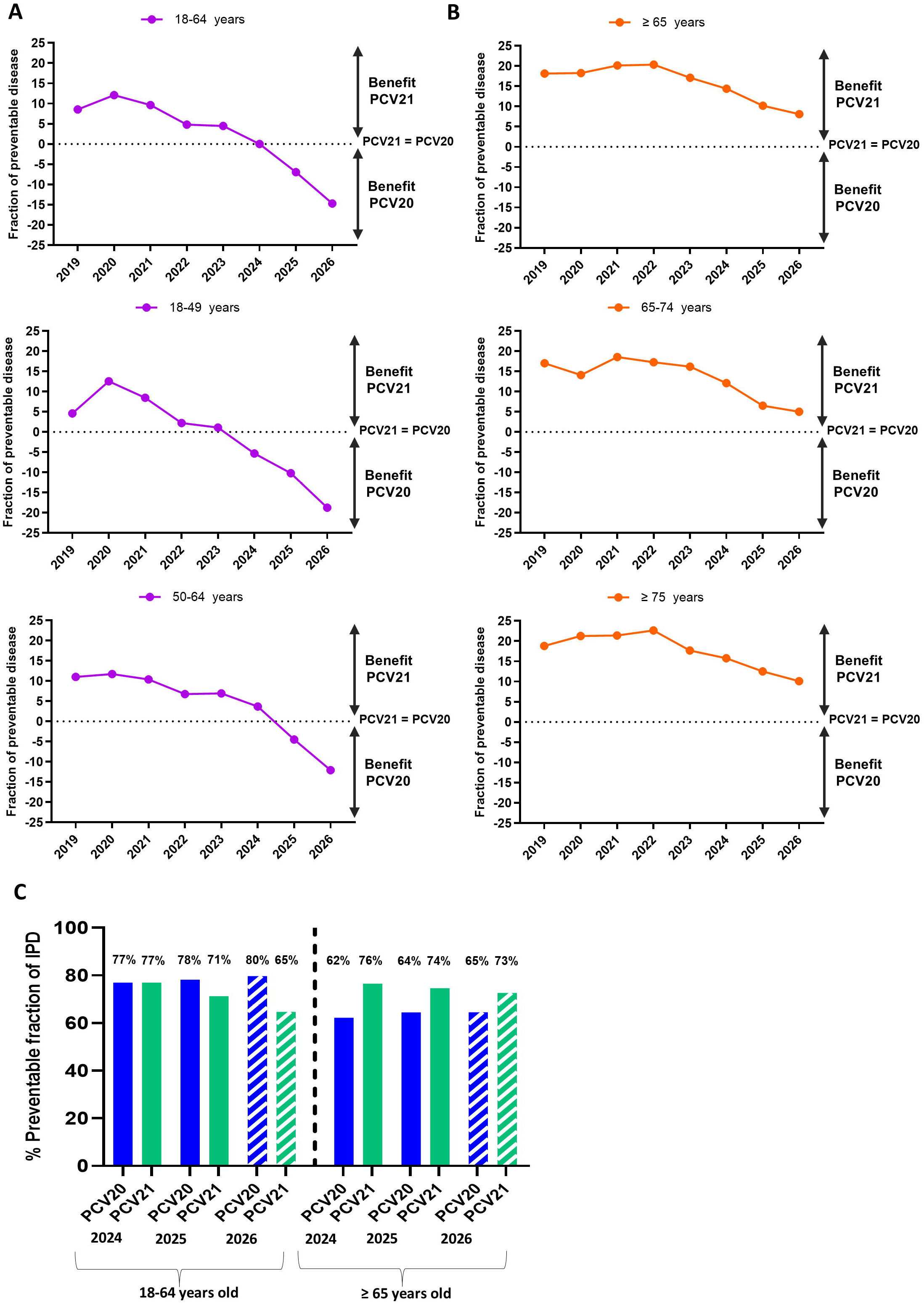
Fraction of disease (%) that can be prevented with PCV20 or PCV21 in adults of different age groups by epidemiological year. Differences in prevention between unique-PCV21 vs unique-PCV20 serotypes in young adults (purple line) (A) and older adults (orange line) (B) during the period 2009-2026. Results were obtained by calculating the difference between the % of IPD cases by unique-PCV21 serotypes minus the % of IPD cases by unique-PCV20 serotypes. Proportion of preventable disease by PCV20 (blue bars) vs PCV21 (green bars) in adults of 18-64 years old and ≥ 65 years old in years 2024 / 2025 (solid bars) and 2026 (hatched bars) (C).

In older adults, the fraction of disease that could be prevented with both vaccines was higher for PCV21 during the study period and for all the age-groups analysed (Figure 5B). However, from 2022 to 2026, we observed a constant reduction in the fraction of disease that could be prevented with PCV21 with a drop from >20% in the year 2022 in benefit of PCV21 to around or less than 10% in 2026 (8% in ≥65 years old, 5% in 65-74 years old and 10% in ≥75 years old) (**Figure 5B**).

Comparison of preventable disease by PCV20 vs PCV21 vaccines, including common PCV20/21 serotypes, between 2025 and 2026 confirmed that in young adults (18-64 years) the differences are increasing in benefit of PCV20 (7% in 2025 vs 15% in 2026). In old adults (≥65 years), however, the difference between both vaccines was less pronounced in 2026 than in previous years (14% in 2024, 10% in 2025 and 8% in just six months of 2026) (**Figure 5C**).

### Distribution of unique-PCV20/PCV21 serotypes causing IPD in adults

The COVID-19 pandemic had a clear impact not only on the burden of IPD in adults but also in the distribution of serotypes causing IPD (**Figure 6**). In young adults (18-64 years old), unique-PCV21 serotypes show a stable trend comparing 2019 vs 2024/2025 with serotype 9N followed by 15A and 24F as the most prevalent (**Figure 6A**). Unique- PCV20 serotypes were less prevalent before the pandemic (year 2019), with a marked increase in incidence in the last two full epidemiological years (2024/2025), especially due to the rise of serotypes 4 and 14 (**Figure 6A**). These findings are of great relevance because with data from 2026, serotypes 4 and 14 are now the third and fourth cause of IPD in young adults, and more surprisingly, serotype 4 is catching serotype 3 that has been the second cause of IPD in this age group in the last years (**Figure 7**).

**Fig 6:**
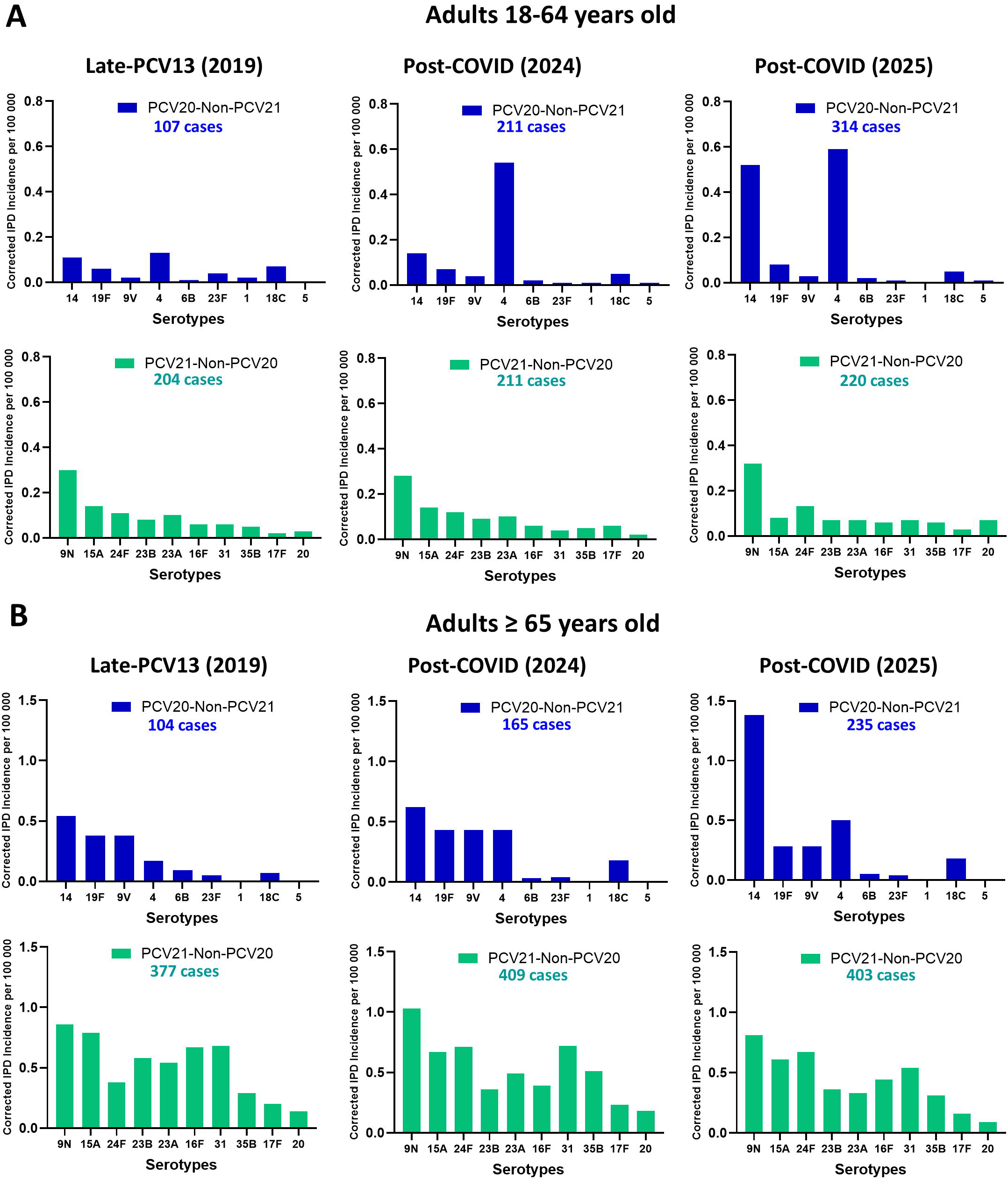
Evolution of IPD incidence by unique-PCV20 vs unique-PCV21 in adults comparing 2019 vs 2024 and 2025. Unique-PCV20 (blue colour) vs unique-PCV21 (green colour). Adults aged 18-64 years (A) and adults aged ≥65 years (B) are represented.

**Fig 7:**
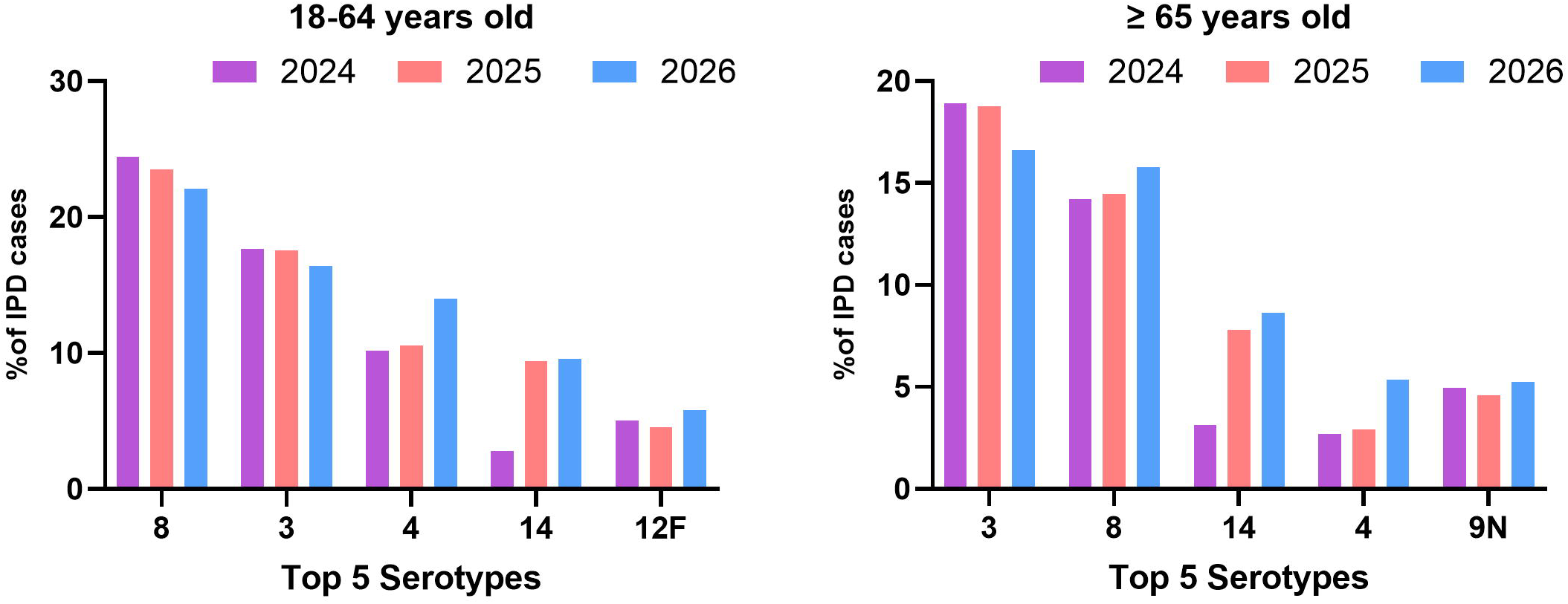
Proportion of IPD cases in adults caused by the five most prevalent serotypes in 2026 in comparison to previous years 2024 and 2025 in young adults (18- 64 years old) and older adults (≥ 65 years old). Year 2024 (purple color), year 2025 (pink color) and year 2026 (blue color).

In older adults (≥ 65 years old), the burden of disease by unique-PCV21 serotypes was higher in the period 2019-2025 compared to unique-PCV20 serotypes, although they show a similar distribution throughout the years without any unusual rise of any unique-PCV21 serotype (**Figure 6B**). Incidence by the most prevalent unique-PCV21 serotype has slightly diminished in the last years. However, among unique-PCV20 serotypes we observed the rise of serotype 4 in the last years and a remarkably high increase of serotype 14 in 2025 (**Figure 6B**). These results are very relevant from the epidemiological perspective because serotype 14 has become the third cause of IPD in adults ≥ 65 years old in the year 2026, and serotype 4 has matched the proportion of cases by serotype 9N, becoming jointly the fourth leading cause of IPD (**Figure 7**).

## Discussion

Evaluation of circulating serotypes causing IPD in adults using the last epidemiological data available is critical to understand the preventive potential and impact on the burden of disease of broader vaccines such as PCV20 and PCV21 that have been approved very recently. Characterization of common serotypes included in both vaccines as well as the evolution of unique serotypes covered by either PCV20 or PCV21 is necessary to address the matter of which vaccine may offer the best coverage in adult population. Moreover, the analysis of non-vaccine serotypes that could be increasing and even the re-emerge of vaccine-covered serotypes is essential to establish the best prophylactic strategies in public health. This 17-year national surveillance study reveals a striking post-pandemic resurgence of serotypes 4 and 14 among adults, reversing the pre-pandemic advantage in theoretical coverage held by PCV21 over PCV20 and narrowing the gap in comparative preventive potential between these two recently approved vaccines. While common PCV20/21 serotypes and unique-PCV20 serotypes declined in the years following paediatric PCV13 introduction — consistent with herd protection — this decline has not persisted, and by 2026 unique-PCV20 serotypes in younger adults have surpassed to unique-PCV21 serotypes for the first time since the pre-vaccine era. This reduction was mainly attributable to herd protection effects induced by paediatric vaccination, given that many of the declining serotypes were also included in PCV13, together with the relatively low vaccination coverage achieved in adults (<30%) (3, 4). This phenomenon of herd protection in adults has been observed in many other countries after the introduction of PCVs in children (19- 21). In contrast, between 2014-2019 we found an increase of common-PCV20/21 serotypes and unique-PCV21 serotypes.

In the case of common-PCV20/21 serotypes, this rise was mainly attributed to serotypes 3 and 8 in adults. Previous studies have confirmed the predominance of hypervirulent lineages (clade Iα/CC180/GPSC12 in serotype 3 and CC53/GPSC3 in serotype 8) containing mutations and allelic variations in virulence factors that increased their virulence and host immune evasion potential (10, 12). This is important because clonal expansion of these lineages has been described globally, contributing to the high prevalence of serotypes 3 and 8 as major causes of IPD worldwide (22-24). The rise of serotype 24F as unique-PCV21 serotype in the pre-pandemic period has been described to be associated with a multidrug-resistant and virulent lineage after PCV13 (GPSC10) (25). In addition, other serotypes such as 9N, 15A or 23A that are included in PCV21 but not in PCV20 have shown increasing trends in the pre-pandemic period after PCV13 introduction and are also related to specific lineages (26, 27). During the COVID-19 pandemic, particularly during the first two years (2020-2021) we observed a sharp reduction of overall IPD across all ages in adults affecting vaccine and non-vaccine serotypes. This effect was influenced by the implementation of non- pharmacological interventions (NPIs) that contributed to the lower transmission pattern of respiratory pathogens as has been previously observed worldwide (1, 28, 29).

During the reopening coinciding with the end of NPIs, we observed a rise in IPD by all serotypes reaching similar levels to before the COVID-19 pandemic, which is consistent with the epidemiological scenario observed in other countries (1, 30). However, since 2023/24 we observed a declining trend of common-PCV20/21 serotypes in all age groups that may be related to the paediatric use of PCV15/20 in Spain since 2024 with high coverage rates (>95%). This reduction contrasts with the increase of unique-PCV20 serotypes in all adult age groups, although in this case, it was mainly due to the rise of serotypes 4 and 14, suggesting a lack of herd protection for these two serotypes (18, 31, 32). In serotype 4, one possible explanation may be due to the emergence of ST15063 within GPSC162 lineage, containing a key mutation in LytA that increases the potential to cause disease, and because serotype 4 strains of this lineage display synergistic infection in the presence of cigarette smoke and with H3N2 flu viruses (11). This is worrisome because the rise of serotype 4 in adults after the COVID-19 pandemic has been described in many other countries, and it may follow a generic trend (14, 33-35). In the case of serotype 14, the causes of the sudden reemergence have not been clarified yet. Hence, the data from 2026 reinforce this increasing pattern as serotypes 4 and 14 have become among the top 5 serotypes causing IPD in adults of all ages, which is consistent with the epidemiological trends in other countries (32) although formal trend-segmentation methods could further characterize the precise timing of these inflection points. This upsurge may change the official vaccine recommendations, expanding the use of PCVs among adults aged ≥50 years, and the threat of serotype 4 in the USA has led to the recommendation of the use of PCVs containing serotype 4, such as PCV20 alone or PCV15 in series with PPSV23 (34, 36, 37). Our data in young adults (18-64 years) are consistent with this recommendation because unique-PCV20 serotypes have surpassed the burden of disease by unique-PCV21 serotypes, and the fraction of IPD that can be prevented with PCV20 is up to 15% higher than PCV21 with our most recent epidemiological data (2026).

The incidence of unique-PCV21 serotypes in adults also returned to pre-pandemic levels after the COVID-19 pandemic, although in recent years it has reached a plateau in most age groups. This rise has been observed in other countries, and therefore, the fraction of disease that can be prevented with PCV21 may be higher than PCV20 using epidemiological data of adults ≥65 years (18, 38). However, our analysis evaluating the differences in preventable disease between PCV21 and PCV20 confirms a continuous decline in the last 4 years, from more than 20% in benefit for PCV21 during 2019-2022 to around or less than 10% during 2025/26. Surveillance studies monitoring the evolution in the following years of unique serotypes contained in PCV21 and PCV20 are necessary to evaluate the potential coverage of these vaccines in adults of different age groups to establish the best prophylactic strategy in the future.

Another critical aspect about the epidemiology of unique serotypes contained in both vaccines is related to the rise of multidrug-resistant strains after the COVID-19 pandemic. Among unique serotypes presenting antibiotic resistance included in PCV20 or PCV21, unique PCV20 serotypes are associated with higher levels of antibiotic resistance to different β-lactam antibiotics, being serotype 15A the unique-PCV21 serotype harbouring high MIC levels (39). Serotype 14 followed by 9V, were the most prevalent in Spain, and associated with high MIC values to penicillin and amoxicillin (31, 39). A similar situation has been observed in North America, reporting the rise of serotype 9V causing IPD in adults of all ages in recent years. Indeed, the upsurge of this serotype has become a serious problem in Canada, as serotype 9V was one of the most highly antimicrobial-resistant serotypes characterized in 2023 (26, 40). Another increasing serotype in the last years is serotype 12F, which is present in PCV20 and PCV21 but not in PCV13 or PCV15. In Spain, this serotype has increased in adults ≥65 years from position 22^nd^ in 2023 (19 cases, 1.43%) to 8^th^ position in the first six months of 2026 (30 cases, 3.15%) and among the top 5 serotypes in 18-64 years (3.60% in 2023 vs 5.79% in 2026) (3). This increase concurs with the epidemiological situation observed in other countries reporting the rise of serotype 12F in adults in recent years (27, 32). Unique-PCV21 serotypes such as serotype 24F present lower levels of antibiotic resistance but are very prevalent in IPD, since PCV13 introduction in Spain (3, 6). Serotype 24F, previously associated with lineages susceptible to antibiotics such as GPSC16, changed rapidly after PCV13 introduction to GPSC6 and GPSC10 in France and Spain (25). Both GPSC6 and GPSC10 are associated with resistance to β- lactams and macrolides (25).

The high diversity of serotypes that can potentially cause pneumococcal disease, with up to 107 different serotypes defined based on their distinct capsular structures, serology, and antigenicity (41), plus the introduction of newer and broader PCVs, requires a complex and multifactorial analysis to implement the best preventive strategy against IPD. For this reason, a critical aspect in the prevention of IPD is to achieve high uptake rates to elicit direct protection in vaccinated groups, including the contribution of herd protection to non-vaccinated individuals. Hence, robust surveillance data describing the most recent epidemiological trends and serotype distribution is essential to monitor the rise of non-vaccine serotypes or even the reemergence of vaccine-covered serotypes (2, 20, 21). Potential limitations for this study are based on regional heterogeneity in vaccine rollout across different regions and lack of current vaccine effectiveness studies in Spain.

To summarize, our results confirm that despite a long-term program of paediatric vaccination with high uptake rates using PCV13, the burden of IPD in adults is still of great relevance with the rise of several vaccine serotypes that require further surveillance. Our study highlights the contribution of broader PCVs such as PCV20 and PCV21 to prevent pneumococcal disease in adults and reduce hospitalization costs. These findings are crucial for establishing future vaccine policies and strategies in adults.

## Data Availability

All epidemiological data requests should be submitted to the corresponding authors. Requests will be assessed for scientific rigor before being granted, and a data sharing agreement might be required.

## Notes

### Disclaimer

The findings and conclusions presented here are those of the authors and do not necessarily represent the official position of Instituto de Salud Carlos III (ISCIII).

### Funding source

This work was supported by Ministerio de Ciencia e Innovación (MICINN) [PID2024-161570OB-I00] and funding from ISCIII (grant PI24CIII/00045/PI21CIII/00012 and COOP24CIII/00032). CP-G was supported by a Formación del Profesorado Universitario fellowship from MICINN (FPU21/01798). This study was also partially funded by grants from Merck Sharp Dohme [grant MISP#IISP101638] and from Pfizer (grant 100177543) although both funders had no intervention in the results of this study. All analyses were conducted independently by the authors.

### Declaration of Competing Interest

JY has received grants from MSD-USA (MISP Call), Pfizer and MEIJI. JY has participated in advisory boards organized by GSK, MSD, and Pfizer. JY declares payments of travel expenses and meeting fees from MSD and Pfizer. JS and MD have participated in advisory boards organized by MSD or Pfizer respectively. CA has received grants from MSD-USA (MISP Call), and Pfizer. CA has participated in advisory boards organized by MSD, and Pfizer. All other authors report no potential conflicts.

## Notes

### Author Declarations

All datasets used in this study were de-identified prior to their use and analysis, and do not contain information that allows individual participants to be identified. The data are analyzed in an aggregated and stratified manner and are derived from routine surveillance activities conducted by the Spanish Pneumococcal Reference Laboratory, recently designated as the European Union Reference Laboratory (EURL) for Public Health on Invasive Bacteria.

## REFERENCES

1. Shaw D, Abad R, Amin-Chowdhury Z, Bautista A, Bennett D, Broughton K, et al. Trends in invasive bacterial diseases during the first 2 years of the COVID-19pandemic: analyses of prospective surveillance data from 30 countries and territories inthe IRIS Consortium. Lancet Digit Health. 2023.

2. Shaw D, Torreblanca RA, Amin-Chowdhury Z, Bautista A, Bennett D,Broughton K, et al. The importance of microbiology reference laboratories and adequatefunding for infectious disease surveillance. Lancet Digit Health. 2024.

3. Perez-Garcia C, Sempere J, de Miguel S, Hita S, Ubeda A, Vidal EJ, et al.Surveillance of invasive pneumococcal disease in Spain exploring the impact of theCOVID-19 pandemic (2019-2023). J Infect. 2024;89(2):106204.

4. de Miguel S, Domenech M, Gonzalez-Camacho F, Sempere J, Vicioso D, Sanz JC, et al.Nationwide Trends of Invasive Pneumococcal Disease in Spain From 2009 Through 2019 in Children and Adults During the Pneumococcal Conjugate VaccineEra. Clin Infect Dis. 2021;73(11):e3778–e87.

5. Dominguez A, Ciruela P, Hernandez S, Garcia-Garcia JJ, Soldevila N, Izquierdo C, et al.Effectiveness of the 13-valent pneumococcal conjugate vaccine in preventinginvasive pneumococcal disease in children aged 7-59 months. A matched case-controlstudy. PLoS One. 2017;12(8):e0183191.

6. Sempere J, Llamosi M, Lopez Ruiz B, Del Rio I, Perez-Garcia C, Lago D, et al.Effect of pneumococcal conjugate vaccines and SARS-CoV-2 on antimicrobialresistance and the emergence of Streptococcus pneumoniae serotypes with reducedsusceptibility in Spain, 2004-20: a national surveillance study. Lancet Microbe.2022;3(10):e744–e52.

7. Garcia-Carretero R, Gil-Prieto R, Hernandez-Barrera V, Gil-de-Miguel A.Epidemiological and clinical impact of pneumococcal disease in Spain in 2023: Anationwide retrospective analysis. Hum Vaccin Immunother. 2025;21(1):2579385.

8. Gil-Prieto R, Allouch N, Jimeno I, Hernandez-Barrera V, Arguedas-Sanz R, Gil-de-Miguel A. Burden of Hospitalizations Related to Pneumococcal Infection in Spain(2016-2020). Antibiotics (Basel). 2023;12(1).

9. Aguinagalde L, Corsini B, Domenech A, Domenech M, Camara J, Ardanuy C,et al. Emergence of Amoxicillin-Resistant Variants of Spain9V-ST156 PneumococciExpressing Serotype 11A Correlates with Their Ability to Evade the Host ImmuneResponse. PLoS One. 2015;10(9):e0137565.

10. Perez-Garcia C, Gonzalez-Diaz A, Domenech M, Llamosi M, Ubeda A, Sanz JC, et al.The rise of serotype 8 is associated with lineages and mutations in the capsularoperon with different potential to produce invasive pneumococcal disease. EmergMicrobes Infect. 2025;14(1):2521845.

11. Perez-Garcia C, Llorente J, Alastuey MEA, Llamosi M, Gil-Prieto R, Laghlali G, et al.Lineage dynamics and risk factors underlying serotype 4 invasivepneumococcal disease in Spain. J Infect Dis. 2026.

12. Perez-Garcia C, Sempere J, Llamosi M, Gonzalez-Diaz A, de Miguel S, Vidal-Alcantara EJ, et al.Upsurge of pneumococcal clade I-alpha/CC180 serotype 3 and itsassociation with a LytA mutation linked to immune evasion and disease potential: anobservational and experimental study. Lancet Microbe. 2026;7(7):101365.

13. Gomez Rial J, Redondo E, Rivero-Calle I, Mascaros E, Ocana D, Jimeno I, et al.Immunofitness in the elderly: The role of vaccination in promoting healthy aging. HumVaccin Immunother. 2026;22(1):2624234.

14. Bertran M, D’Aeth JC, Abdullahi F, Eletu S, Andrews NJ, Ramsay ME, et al.Invasive pneumococcal disease 3 years after introduction of a reduced 1 + 1 infant 13-valent pneumococcal conjugate vaccine immunisation schedule in England: aprospective national observational surveillance study. Lancet Infect Dis. 2024.

15. Nikhab A, Patel T, Rooney G, Wasti S, Abdullahi F, D’Aeth JC, et al.Assessingthe potential impact of the 20-valent (PCV20) and an adult 21-valent (aPCV21)pneumococcal conjugate vaccine on invasive pneumococcal disease in England.Vaccine. 2026;75:128246.

16. Hao L, Kuttel MM, Ravenscroft N, Thompson A, Prasad AK, Gangolli S, et al.Streptococcus pneumoniae serotype 15B polysaccharide conjugate elicits a cross-functional immune response against serotype 15C but not 15A. Vaccine.2022;40(33):4872–80.

17. De Wals P. A new approach to define the optimal immunization strategy againstpneumococcal disease: the example of Canada. Epidemiol Infect. 2025;153:e55.

18. Rieger R, Yahav D, Margalit I, Wieder-Finesod A, Azulay H, Ghanem-Zoubi N,et al. Anticipating impact of implementing PCV20 or PCV21 vaccines for older adultsin the immunisation programme for invasive pneumococcal disease, using nationwidesurveillance data, Israel, 2009 to 2024. Euro Surveill. 2026;31(15).

19. Yang Y, Knoll MD, Herbert C, Bennett JC, Feikin DR, Garcia Quesada M, et al.Global impact of 10- and 13-valent pneumococcal conjugate vaccines on pneumococcalmeningitis in all ages: The PSERENADE project. J Infect. 2025;90(3):106426.

20. Bennett JC, Deloria Knoll M, Kagucia EW, Garcia Quesada M, Zeger SL,Hetrich MK, et al.Global impact of ten-valent and 13-valent pneumococcal conjugatevaccines on invasive pneumococcal disease in all ages (the PSERENADE project): aglobal surveillance analysis. Lancet Infect Dis. 2025;25(4):457–70.

21. Garcia Quesada M, Peterson ME, Bennett JC, Hayford K, Zeger SL, Yang Y, et al.Serotype distribution of remaining invasive pneumococcal disease after extensiveuse of ten-valent and 13-valent pneumococcal conjugate vaccines (the PSERENADEproject): a global surveillance analysis. Lancet Infect Dis. 2025;25(4):445–56.

22. Azarian T, Mitchell PK, Georgieva M, Thompson CM, Ghouila A, Pollard AJ,et al. Global emergence and population dynamics of divergent serotype 3 CC180 pneumococci. PLoS Pathog. 2018;14(11):e1007438.

23. Lekhuleni C, Ndlangisa K, Sanches Ferreira AD, Cheng HR, Lorenz O,Kleynhans J, et al.Clonal expansion of global pneumococcal sequence cluster 3 withinserotype 8 after 13-valent pneumococcal conjugate vaccine introduction, South Africa.Microb Genom. 2026;12(6).

24. D’Aeth JC, Bertran M, Abdullahi F, Eletu S, Hani E, Fry NK, et al.Whole-genome sequencing, strain composition, and predicted antimicrobial resistance ofStreptococcus pneumoniae causing invasive disease in England in 2017-20: aprospective national surveillance study. Lancet Microbe. 2025;6(7):101102.

25. Lo SW, Mellor K, Cohen R, Alonso AR, Belman S, Kumar N, et al.Emergenceof a multidrug-resistant and virulent Streptococcus pneumoniae lineage mediatesserotype replacement after PCV13: an international whole-genome sequencing study.Lancet Microbe. 2022;3(10):e735–e43.

26. Golden A, Griffith A, Lefebvre B, McGeer A, Tyrrell G, Kus J, et al.Invasivepneumococcal disease surveillance in Canada, 2023. Can Commun Dis Rep. 2026;52(1-2):36–46.

27. Ndlangisa KM, Lekhuleni C, Skosana H, de Gouveia L, Meiring S, Walaza S, et al.Population snapshot of Streptococcus pneumoniae causing invasive disease amongadults aged >/=18 years in South Africa before and after implementation ofpneumococcal conjugate vaccines in 2005-2020. Microb Genom. 2025;11(11).

28. Brueggemann AB, Jansen van Rensburg MJ, Shaw D, McCarthy ND, Jolley KA, Maiden MCJ, et al.Changes in the incidence of invasive disease due toStreptococcus pneumoniae, Haemophilus influenzae, and Neisseria meningitidis duringthe COVID-19 pandemic in 26 countries and territories in the Invasive RespiratoryInfection Surveillance Initiative: a prospective analysis of surveillance data. LancetDigit Health. 2021;3(6):e360–e70.

29. Prasad N, Rhodes J, Deng L, McCarthy NL, Moline HL, Baggs J, et al.Changesin the Incidence of Invasive Bacterial Disease During the COVID-19 Pandemic in theUnited States, 2014-2020. J Infect Dis. 2023;227(7):907–16.

30. Perniciaro S, van der Linden M, Weinberger DM. Reemergence of InvasivePneumococcal Disease in Germany During the Spring and Summer of 2021. Clin InfectDis. 2022;75(7):1149–53.

31. Sanz-Moreno JC, Garrido-Estepa M, Inigo-Martinez J, Martin-Martinez F,Humanes-Navarro AM, Garrido-Buenache A, et al.Increase in serotypes 4 and 14 ininvasive pneumococcal disease in adults aged 18 to 59 years. A retrospectivepopulation-based descriptive study during the period 2007-2025 in the Community ofMadrid, Spain. European journal of clinical microbiology & infectious diseases :official publication of the European Society of Clinical Microbiology. 2026.

32. Cuypers L, Dambre C, Desmet S. Exceptional high number of IPD cases inwinter season 2024-2025 in Belgium in concomitance with rise in vaccine serotypes.Vaccine. 2025;64:127763.

33. Manca MF, Silvola J, Czarnecki J, Sequeira Neto J, Kanerva M, Kaukavuori H,et al. Third Streptococcus pneumoniae disease outbreak involving serotype 4-ST801 ina shipyard, Finland, May to June 2025. Euro Surveill. 2025;30(41).

34. Beall B, Walker H, Tran T, Li Z, Varghese J, McGee L, et al.Upsurge ofConjugate Vaccine Serotype 4 Invasive Pneumococcal Disease Clusters Among AdultsExperiencing Homelessness in California, Colorado, and New Mexico. J Infect Dis.2021;223(7):1241–9.

35. Cuypers L, Sanchez GJ, Herrera Avila JP, Naesens R, Antoine-Moussiaux T,Herens S, et al.Rapid increase in serotype 4 invasive pneumococcal disease amongvulnerable young male adults in Belgium (2020-2024). Int J Infect Dis.2026;163:108216.

36. Sutcliffe CG, Sergent VM, Yazzie D, Brasinikas G, Christensen L, Clark M, et al.Post-COVID rebound in invasive pneumococcal disease driven by resurgence ofserotype 4 among American Indian individuals in the Southwest United States. J InfectDis. 2026.

37. Kobayashi M, Leidner AJ, Gierke R, Xing W, Accorsi E, Moro P, et al.Expanded Recommendations for Use of Pneumococcal Conjugate Vaccines AmongAdults Aged >/=50 Years: Recommendations of the Advisory Committee on Immunization Practices -United States, 2024. MMWR Morb Mortal Wkly Rep.2025;74(1):1–8.

38. Martino A, Urrego-Reyes J, Cervenka J, Paluru MK, Cossrow N. Invasivepneumococcal disease in Latin America: serotype distribution and vaccine coverageamong adults in Mexico, Brazil, and Argentina. Vaccine. 2026;88:128947.

39. Llamosi M, Perez-Garcia C, Ubeda A, Vidal-Alcantara EJ, Pareja-Cerban I,Gomez-Rubio E, et al.Antimicrobial activity of beta-lactam antibiotics againstpneumococcal isolates causing pneumococcal disease in adults immediately before andafter the COVID-19 pandemic in Spain (2019-2020). Front Pharmacol.2025;16:1658431.

40. Griffith A, Golden AR, Lefebvre B, McGeer A, Tyrrell GJ, Zhanel GG, et al.Invasive pneumococcal disease surveillance in Canada, 2021-2022. Can Commun DisRep. 2024;50(5):121–34.

41. Lorenz O, King AC, Hung HCH, Ganaie FA, Wyllie AL, Manna S, et al.SeroBA(v2.0) and SeroBAnk: a robust genome-based serotyping scheme andcomprehensive atlas of capsular diversity in Streptococcus pneumoniae. MicrobGenom. 2025;11(10).

