## Supplementary Table S1 for "National trends of unique-PCV20 / PCV21 serotypes, common PCV20-PCV21 and non-vaccine serotypes causing invasive pneumococcal disease in adults in Spain during the period 2009-2026"

**Supplementary Table S1**. Number of cases, corrected incidence of invasive pneumococcal disease and incidence rate ratios in the last full epidemiological year comparing pre-PCV13 introduction (2009), pre-COVID-19 pandemic (2019) and last full epidemiological year (2025). IRR, incidence rate ratio

|  | **2009** | | **2019** | | **2025** | | **IRR 2025 vs 2019** | **95% CI** | **IRR 2025 vs 2009** | **95% CI** | **IRR 2019 vs 2009** | **95% CI** |
| --- | --- | --- | --- | --- | --- | --- | --- | --- | --- | --- | --- | --- |
|  | cases | incidence (per 100000) | cases | incidence (per 100000) | cases | incidence (per 100000) |  |  |  |  |  |  |
| **18 to 64 years old** | 1278 | 5.26 | 1135 | 4.75 | 1358 | 5.48 | 1.15 | 1.06-1.24 | 1.04 | 0.96-1.12 | 0.9 | 0.83-0.97 |
| COMMON PCV20-21 | 673 | 2.77 | 761 | 3.19 | 748 | 3.02 | 0.95 | 0.86-1.05 | 1.09 | 0.98-1.21 | 1.15 | 1.04-1.28 |
| PCV20-NON21 | 408 | 1.68 | 107 | 0.45 | 314 | 1.27 | 2.83 | 2.27-3.52 | 0.75 | 0.65-0.87 | 0.27 | 0.22-0.33 |
| PCV21-NON-20 | 146 | 0.60 | 204 | 0.85 | 220 | 0.89 | 1.04 | 0.86-1.26 | 1.48 | 1.20-1.82 | 1.42 | 1.15-1.76 |
| NON-VAC | 51 | 0.21 | 63 | 0.26 | 76 | 0.31 | 1.16 | 0.83-1.62 | 1.46 | 1.02-2.08 | 1.26 | 0.87-1.82 |
| **18 to 49 years old** | 765 | 4.25 | 436 | 2.74 | 579 | 3.61 | 1.32 | 1.17-1.49 | 0.85 | 0.76-0.95 | 0.64 | 0.57-0.72 |
| COMMON PCV20-21 | 368 | 2.05 | 301 | 1.89 | 327 | 2.04 | 1.08 | 0.92-1.26 | 1 | 0.86-1.16 | 0.93 | 0.80-1.08 |
| PCV20-NON21 | 277 | 1.54 | 47 | 0.30 | 147 | 0.92 | 3.1 | 2.23-4.31 | 0.6 | 0.49-0.73 | 0.19 | 0.14-0.26 |
| PCV21-NON-20 | 91 | 0.51 | 67 | 0.42 | 88 | 0.55 | 1.3 | 0.95-1.79 | 1.09 | 0.81-1.46 | 0.83 | 0.61-1.14 |
| NON-VAC | 29 | 0.16 | 21 | 0.13 | 17 | 0.11 | 0.8 | 0.42-1.52 | 0.66 | 0.36-1.20 | 0.82 | 0.47-1.44 |
| **50 to 64 years old** | 513 | 8.12 | 699 | 9.00 | 779 | 8.90 | 0.99 | 0.89-1.10 | 1.1 | 0.98-1.23 | 1.11 | 0.99-1.24 |
| COMMON PCV20-21 | 305 | 4.83 | 460 | 5.93 | 421 | 4.81 | 0.81 | 0.71-0.92 | 1 | 0.86-1.16 | 1.23 | 1.06-1.42 |
| PCV20-NON21 | 131 | 2.07 | 60 | 0.77 | 167 | 1.91 | 2.47 | 1.84-3.32 | 0.92 | 0.73-1.16 | 0.37 | 0.27-0.50 |
| PCV21-NON-20 | 55 | 0.87 | 137 | 1.76 | 132 | 1.51 | 0.85 | 0.67-1.08 | 1.73 | 1.26-2.37 | 2.03 | 1.48-2.78 |
| NON-VAC | 22 | 0.35 | 42 | 0.54 | 59 | 0.67 | 1.25 | 0.84-1.86 | 1.94 | 1.19-3.17 | 1.55 | 0.93-2.60 |
| **65 to 74 years old** | 426 | 14.10 | 564 | 15.41 | 644 | 15.66 | 1.02 | 0.91-1.14 | 1.11 | 0.98-1.25 | 1.09 | 0.96-1.24 |
| COMMON PCV20-21 | 258 | 8.54 | 348 | 9.51 | 367 | 8.93 | 0.94 | 0.81-1.09 | 1.04 | 0.89-1.22 | 1.11 | 0.94-1.30 |
| PCV20-NON21 | 94 | 3.11 | 35 | 0.96 | 89 | 2.16 | 2.26 | 1.53-3.34 | 0.7 | 0.52-0.94 | 0.31 | 0.21-0.46 |
| PCV21-NON-20 | 56 | 1.85 | 131 | 3.58 | 131 | 3.19 | 0.89 | 0.70-1.13 | 1.72 | 1.26-2.35 | 1.93 | 1.41-2.64 |
| NON-VAC | 18 | 0.60 | 50 | 1.37 | 57 | 1.39 | 1.01 | 0.69-1.48 | 2.33 | 1.37-3.96 | 2.29 | 1.34-3.92 |
| **75 to 84 years old** | 756 | 32.31 | 940 | 39.50 | 1008 | 36.29 | 0.92 | 0.84-1.01 | 1.12 | 1.02-1.23 | 1.22 | 1.11-1.34 |
| COMMON PCV20-21 | 420 | 17.95 | 529 | 22.23 | 463 | 16.67 | 0.75 | 0.66-0.85 | 0.93 | 0.81-1.06 | 1.24 | 1.09-1.41 |
| PCV20-NON21 | 164 | 7.01 | 69 | 2.90 | 146 | 5.26 | 1.81 | 1.36-2.41 | 0.75 | 0.60-0.94 | 0.41 | 0.31-0.54 |
| PCV21-NON-20 | 133 | 5.68 | 246 | 10.34 | 272 | 9.79 | 0.95 | 0.80-1.13 | 1.72 | 1.40-2.12 | 1.82 | 1.47-2.25 |
| NON-VAC | 39 | 1.67 | 96 | 4.03 | 127 | 4.57 | 1.13 | 0.87-1.47 | 2.74 | 1.91-3.92 | 2.42 | 1.67-3.51 |
| **≥65 years old** | 1182 | 19.29 | 1504 | 20.76 | 1652 | 20.29 | 0.98 | 0.91-1.05 | 1.05 | 0.97-1.13 | 1.08 | 1.00-1.17 |
| COMMON PCV20-21 | 678 | 11.07 | 877 | 12.10 | 830 | 10.19 | 0.84 | 0.76-0.92 | 0.92 | 0.83-1.02 | 1.09 | 0.99-1.20 |
| PCV20-NON21 | 258 | 4.21 | 104 | 1.44 | 235 | 2.89 | 2.01 | 1.60-2.53 | 0.69 | 0.58-0.82 | 0.34 | 0.27-0.43 |
| PCV21-NON-20 | 189 | 3.09 | 377 | 5.20 | 403 | 4.95 | 0.95 | 0.83-1.09 | 1.6 | 1.35-1.90 | 1.69 | 1.42-2.01 |
| NON-VAC | 57 | 0.93 | 146 | 2.02 | 184 | 2.26 | 1.12 | 0.90-1.39 | 2.43 | 1.81-3.27 | 2.17 | 1.60-2.95 |
| **≥85 years old** | 290 | 37.85 | 457 | 37.90 | 466 | 37.18 | 0.98 | 0.86-1.11 | 0.98 | 0.85-1.13 | 1 | 0.86-1.16 |
| COMMON PCV20-21 | 165 | 21.54 | 253 | 20.98 | 195 | 15.56 | 0.74 | 0.61-0.89 | 0.72 | 0.59-0.89 | 0.97 | 0.80-1.18 |
| PCV20-NON21 | 64 | 8.35 | 34 | 2.82 | 73 | 5.82 | 2.07 | 1.38-3.11 | 0.7 | 0.50-0.98 | 0.34 | 0.22-0.52 |
| PCV21-NON-20 | 48 | 6.27 | 110 | 9.12 | 129 | 10.29 | 1.13 | 0.88-1.46 | 1.64 | 1.18-2.28 | 1.46 | 1.04-2.05 |
| NON-VAC | 13 | 1.70 | 60 | 4.98 | 69 | 5.50 | 1.11 | 0.79-1.57 | 3.24 | 1.79-5.86 | 2.93 | 1.61-5.34 |
| **TOTAL** | **5210** | **121.18** | **5735** | **130.06** | **6486** | **127.40** | **0.98** | **0.95-1.02** | **1.05** | **1.01-1.09** | **1.07** | **1.03-1.11** |
